# THE COST-EFFECTIVENESS OF DEPRESSION SCREENING FOR THE GENERAL ADULT POPULATION

**DOI:** 10.1101/2021.05.07.21256842

**Authors:** Melike Yildirim, Bradley N Gaynes, Pinar Keskinocak, Brian W Pence, Julie Swann

**Author notes:** **Corresponding Author:** Julie Swann, Ph.D., Department of Industrial and Systems Engineering, North Carolina State University, Raleigh, North Carolina, USA, 111 Lampe Drive CB7906, Raleigh, NC 27695.; e□mail.

## Abstract

**Background:** Depression is a treatable disease, and untreated depression can lead to serious health complications and decrease the quality of life. Therefore, prevention, early identification, and treatment efforts are essential. Screening has an essential role in preventive medicine in the general population. Ideally, screening tools detect patients early enough to manage the disease and reduce symptoms. We aimed to determine the cost-effectiveness of routine screening schedules.

**Methods:** We used a discrete-time nonstationary Markov model to simulate the progression of depression. We used Monte Carlo techniques to simulate the stochastic model for 20 years or during the lifetime of individuals. Baseline and screening scenario models with screening frequencies of annual, 2-year, and 5-year strategies were compared based on incremental cost-effectiveness ratios (ICER). Monte Carlo (MC) simulation and one-way sensitivity analysis were conducted to manage uncertainties.

**Results:** In the general population, all screening strategies were cost-effective compared to the baseline. However, male and female populations differed based on cost over quality-adjusted life years (QALY). Females had lower ICERs, and annual screening had the highest ICER for females, with 11,134$/QALY gained. In contrast, males had around three times higher ICER, with annual screening costs of 34,065$/QALY gained.

**Limitations:** We assumed that the screening frequency was not changing at any time during the screening scenario. In our calculations, false-positive cases were not taking into account.

**Conclusions:** Considering the high lifetime prevalence and recurrence rates of depression, detection and prevention efforts can be one critical cornerstone to support required care. Our analysis combined the expected benefits and costs of screening and assessed the effectiveness of screening scenarios. We conclude that routine screening is cost-effective for all age groups of females and young, middle-aged males.

## INTRODUCTION

Depression is one of the most common mental health conditions and a leading cause of disability that results in substantial impairment (Goodwin, 2006). Both major depression and minor depression present with either depressed mood or loss of interest or pleasure in usual activities along with other symptoms, and the signs are present for two weeks or longer. Minor depression, with a few mild symptoms, can be resolved without treatment, but it may evolve into major depression. Major depression with moderate or severe symptoms significantly impacts life quality and may require treatment from a mental health specialist. However, most depression cases are treated in general medical settings in practice (Kessler et al., 2010).

On average, 22.9% of females and 15.1% of males experience at least one episode of major depression in their lifetime, and 43.3% of patients are not receiving any treatment (Kessler et al., 2010). Between 1999 and 2019, suicide rates, whether with an underlying diagnosis of depression or not, increased by 33% in the overall population (Center for Disease Control and Prevention, 2020). A 50% increase in suicide rates was observed among women from 2000 to 2016 (Center for Disease Control and Prevention, 2020).

Depression co-occurs highly with other physical illnesses, which causes health and economic burdens to individuals and society. It is estimated that depression cost $210 billion in 2010, including direct and indirect costs (e.g., loss of productivity) (Greenberg et al., 2015). Considering high prevalence and low detection rates, improved management of disease may reduce health spending and improve patients ‘ quality and quantity of life. Due to the high cost and the negative impacts on patients ‘ overall health, improved detection efforts may be adopted by policymakers and healthcare agencies.

Routine depression screening can be used to improve the recognition of depressive episodes. In some health systems, screening is commonly used; in other settings, implementation is limited (O ‘Connor et al., 2016). Therefore, in current clinical practice, screening measures are variable. According to the National Ambulatory Medical Care Survey (NAMCS) in the United States, only 2.29 percent of primary care visit patients were screened for depression in 2010 (National Center for Health Statistics, 2018). Whereas, in postpartum populations, screening is implemented on a large scale (79%) (Sleath et al., 2007). The U.S. Preventive Services Task Force (USPSTF) suggests screening of patients with a system that accurately diagnoses, treats, and follows up with the patients. Still, the benefit of screening, optimal screening interval, and timing is unknown (O ‘Connor et al., 2016; U.S. Preventive Services Task Force, 2016). Studies in literature questioned the USPSTF recommendations (Thombs et al., 2012), and most of them are limited to show the cost-effectiveness of routine depression screening (Valenstein et al., 2001).

The frequency of screening of the general population in different health settings is uncertain. Therefore, evaluation of medical benefits (e.g., decrease the recurrence, relapses, improve treatment success and increase the symptom-free days) and costs of routine depression screening strategies remain essential. In this study, we aimed to determine the cost-effectiveness of routine screening schedules. We assessed the various routine screening frequencies for the general population and specific age groups of females and males.

## METHODS

### Model Structure

A natural history model was introduced where probabilities were assigned to each health state based on patient characteristics such as age, gender, and history of depression. We used a discrete-time nonstationary Markov chain model, a probability distribution that changes as health state, history of depression changes, and time progress. The model consists of ten health states: healthy, major depression without treatment, major depression with treatment, partial remission, full remission from major depression, minor depression without treatment, minor depression with treatment, full remission from minor depression, suicide, and death from other causes (Figure 1). States were similar to Valenstein et al. ‘s and others ‘ (Ross et al., 2019; Valenstein et al., 2001) model.

**Figure 1.**
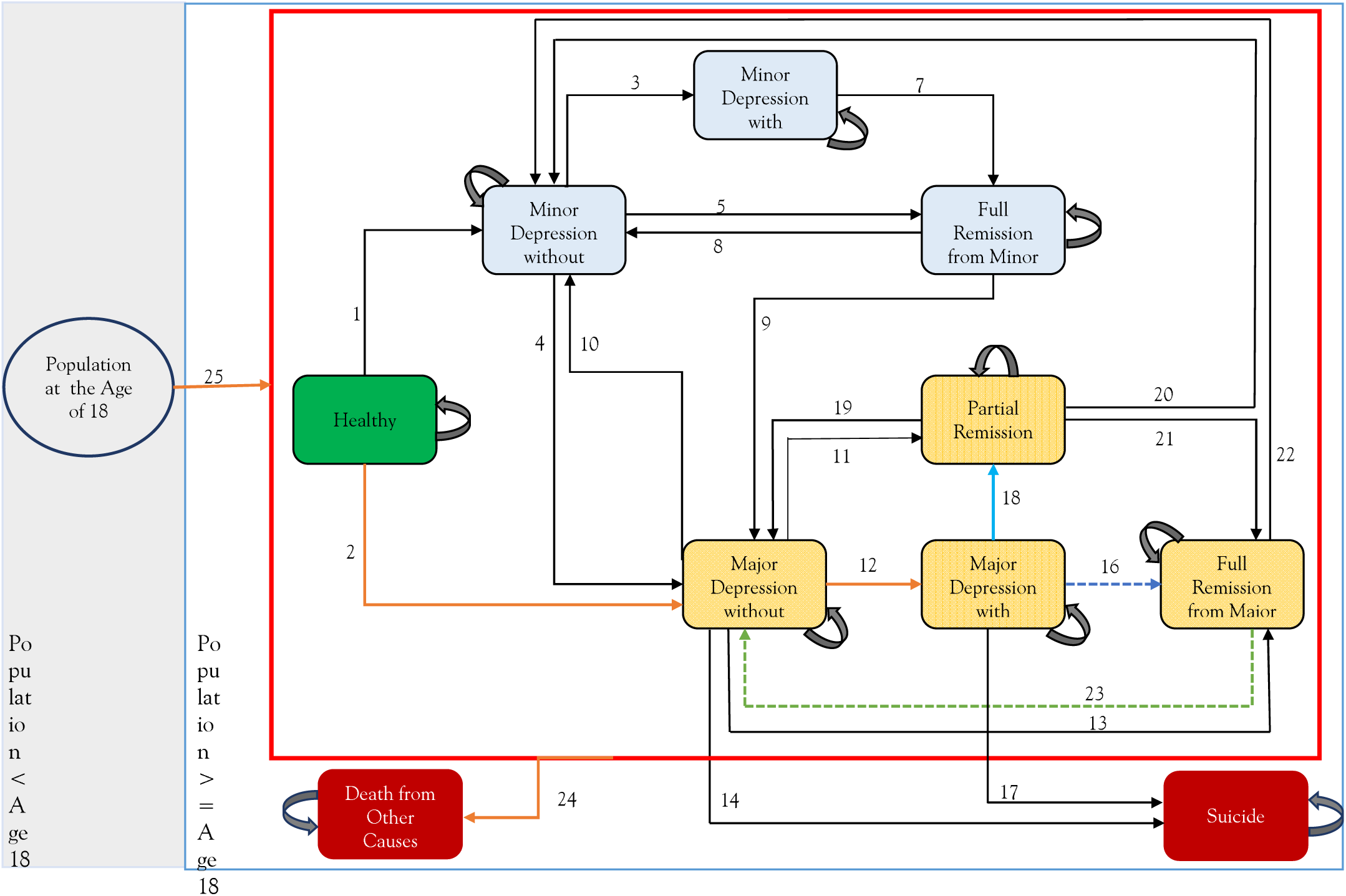
Markov Health State Model

We classified the existence of 2 to 4 depressive symptoms (e.g., feelings of worthlessness, loss of interest, sleep disturbance or sleeping too much, reduced appetite, fatigue) as minor depression (Fils et al., 2010). We considered patients in partial remission of current major depression when they have experienced the residual symptoms of the most recent episode of depression (National Institute for Health and Clinical Excellence: Guidance, 2010). Healthy state included individuals who were never depressed (minor or major).

We considered the following scenarii i) baseline (no screening), ii) annual screening, iii) screening every 2 years, and iv) screening every 5 years. We evaluated the case of screening for the overall population, starting at the age of 18. For screening scenarii, we assumed that the screening tool could increase the probability of patients to diagnosis. We used Monte Carlo techniques to simulate the stochastic model for 20 years or during the lifetime of individuals. Since depression treatment generates long-term benefits, we modeled for a 20-year time horizon, similar to others in the literature (Mark T. Linthicum et al., 2016; Valenstein et al., 2001).

We ran each scenario for 1000 replications. At the entry, patients were assigned their initial states based on prevalence, and they transitioned between states at the end of each year. The initial population of 100,000 individuals was introduced to the system based on U.S. age distribution (Howden and Meyer, 2010) and a constant number of people preserved in the system. In each time unit, the number of 18-year-old individuals entering the system was equal to the number of people leaving the system from states of suicide or death from other causes. We used R (version 3.6.2) in our analysis.

### Model Parameters

We derived the model parameters from published data. Time-sensitive transition probabilities were used, where the mortality probabilities were increased by age. We used annual transitions that depend on patient histories, such as the number of previous episodes, treatment status, time spent without treatment, and demographics (age, gender) if applicable. Table 1 shows the parameters that were used in the baseline model and their sources.

**Table 1.**
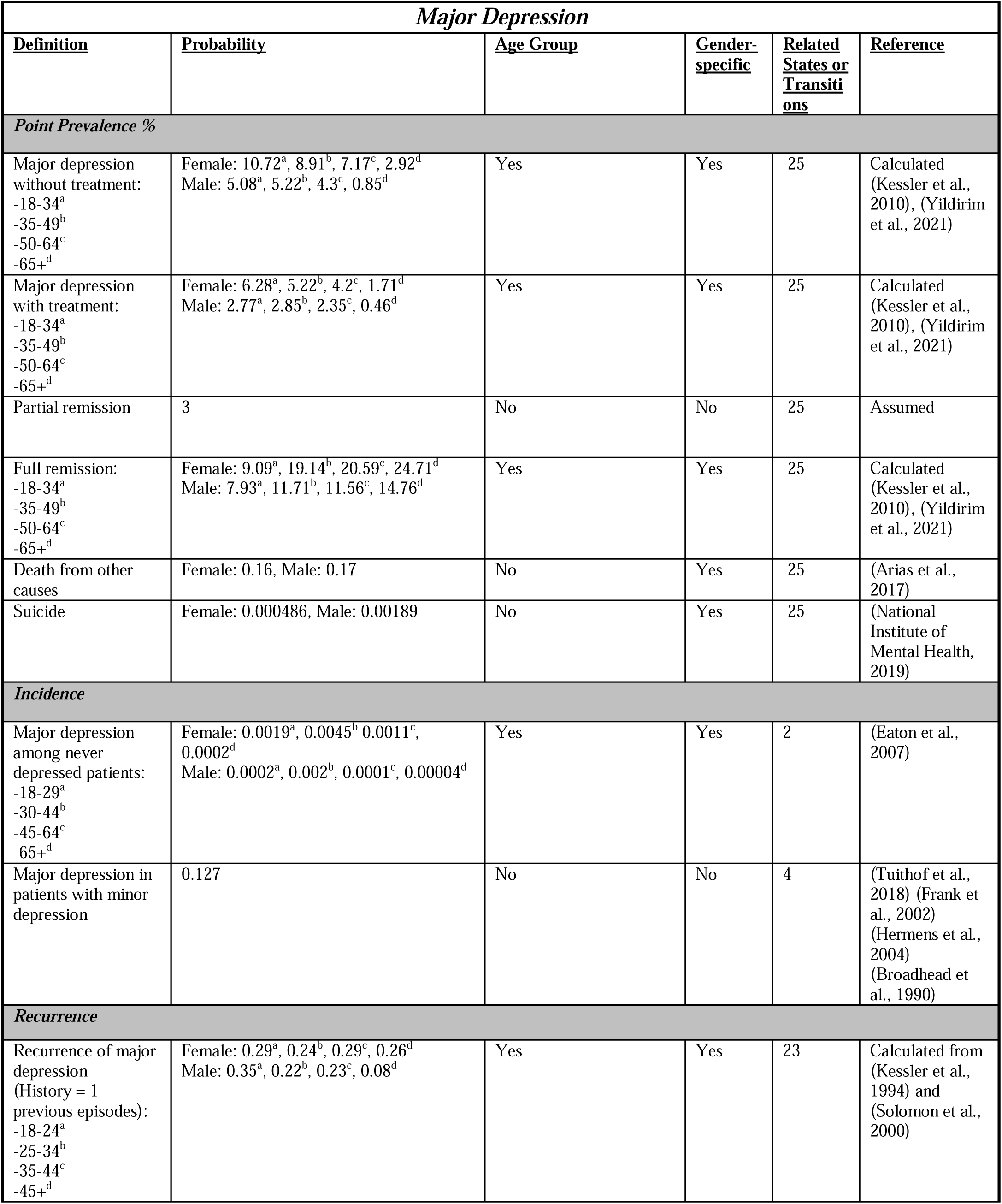
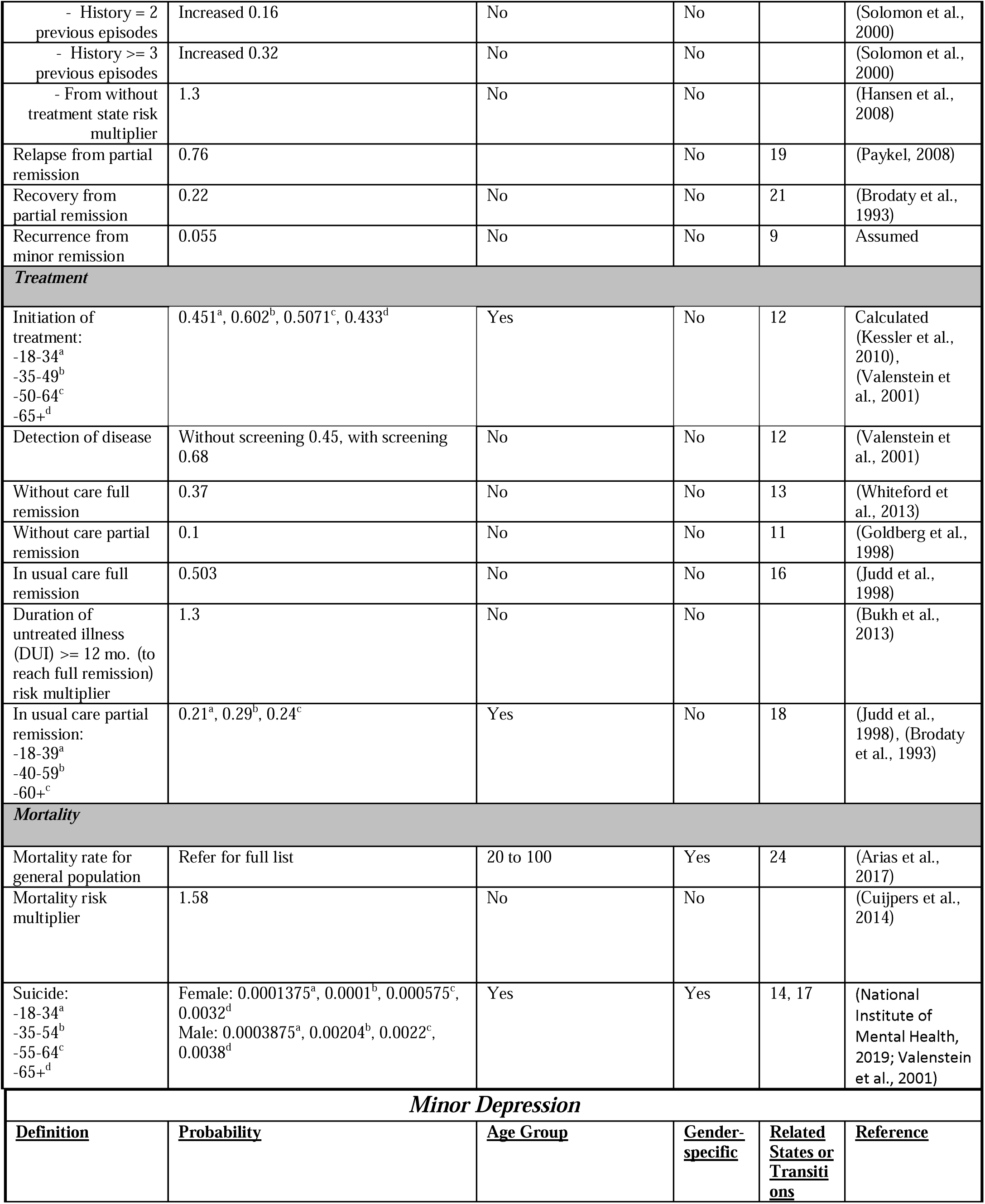
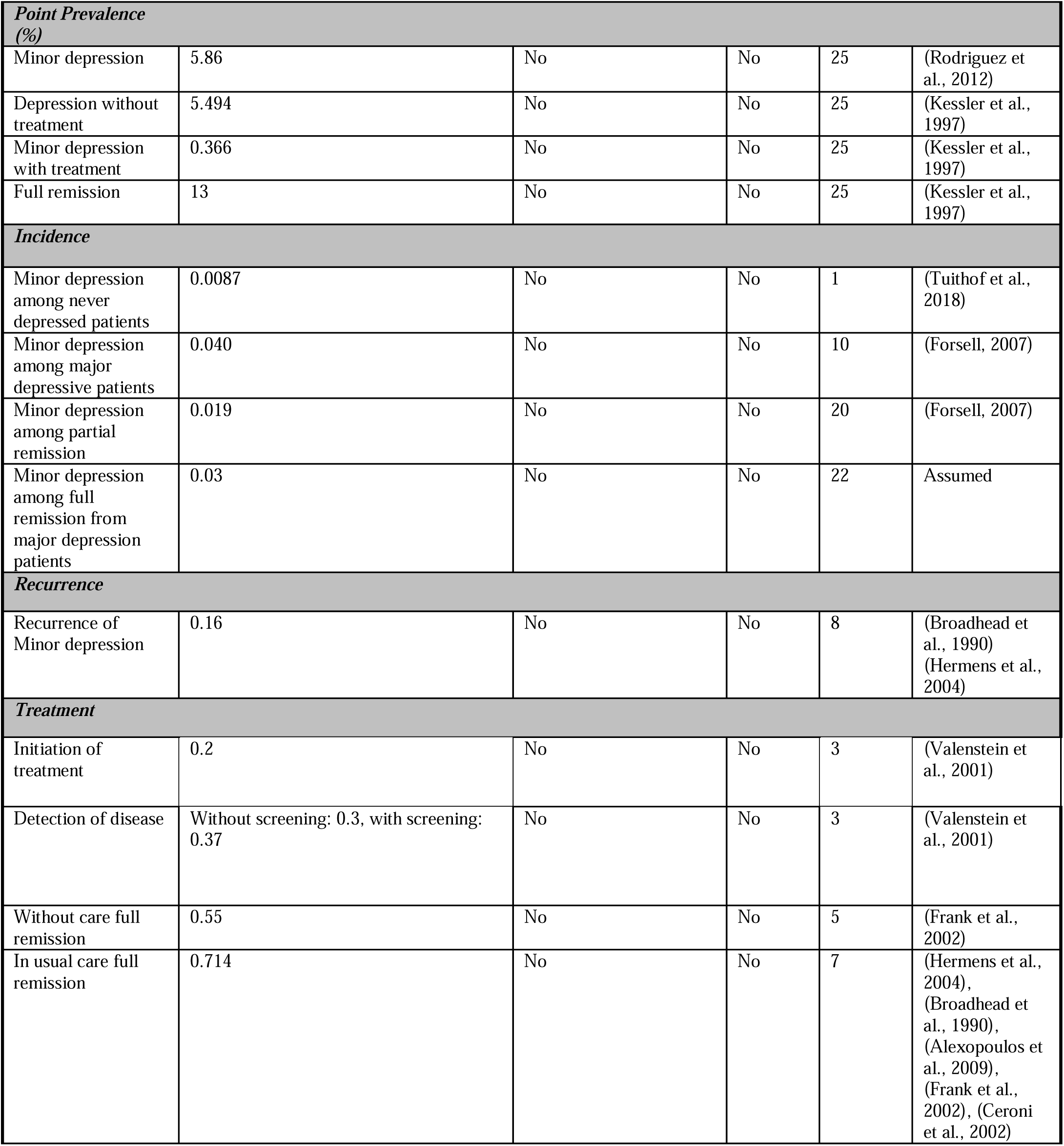
Major and minor depression parameters

When there was no consensus in the literature for a parameter value, we chose conservative estimates. For example, for the incidence of major depression, we used the lower bound of rates reported in the ECA study (Eaton et al., 2007). Consequently, our results were not biased towards screening.

#### Prevalence and Incidence

The ECA study ‘s incidence rates yield a 50% lifetime prevalence of major depression (Eaton et al., 1997), which is higher than reported in the literature. There are arguments around the potential discordance between incidence and lifetime prevalence rates of major depression (Takayanagi et al., 2014).

To be consistent with the reported incidence rates from national surveys, we used a lower bound of incidence (Eaton et al., 2007). We integrated the matching lifetime prevalence using a recall bias of 31.9% for females and 16.3% for males, and a 12-month prevalence recall bias of 25.5% for females and 9% for males, which were obtained from the literature (Yildirim et al., 2021).

#### Detection and treatment

We had a detection rate of 45% for major depression and 30% for minor depression in the baseline model (Valenstein et al., 2001). 43 to 60% (Kessler et al., 2010) of patients who initiated the treatment from major depression depend on the age of onset, whereas 20% (Valenstein et al., 2001) of cases with minor depression received treatment. The multiplication of these two probabilities ((detection rate)*(initiation of treatment)) represented the transitions from without treatment to with treatment state.

Treatment increased the full remission rate by 35% (0.37 without treatment vs. 0.50 with treatment) for major depression and 29% (0.55 without treatment vs. 0.71 with treatment) for minor depression (Table 1).

Early treatment efforts had an impact on the outcome of antidepressant treatment (Kraus et al., 2019). Duration of untreated illness (DUI) longer than 6 months decreased the effect of antidepressant treatment. Patients who spent more than 12 months without treatment status have reduced remission likelihood by one-third (Bukh et al., 2013).

Transition to remission state depended on the healthcare provider and the type of treatment. We assumed that 31% of major depression and 25% of minor depression cases were treated by a specialist (Valenstein et al., 2001). Patients were treated with medications in primary care (Valenstein et al., 2001). Whereas psychotherapy, medication, or a combination of these two was used during the treatment of a specialist.

#### Relapse and recurrence

Major depression is a recurrent mental health condition; around 67% of patients have at least one recurrence every 10 years (Solomon et al., 2000). Patients with residual symptoms were 3 times more likely to relapse than the fully recovered patients (0.76 from partial remission vs. 0.25 from full remission) (Judd et al., 1998). The number of previous episodes and treatment status affected the probability of recurrences. We obtained base recurrence rates for patients who had a history of cases pertaining to depression from (Solomon et al., 2000) and patients who had one previous case and determined that they indicated an increased risk of recurrence by 16% compared to the base. Patients with 2 or more previous episodes had an additional 32% risk of recurrence (Solomon et al., 2000).

Patients who recovered from major remission without appropriate medication or other treatments were more likely (relative risk of 1.52 to 2.69 (Gelenberg et al., 2003; Lustman et al., 2006; Montgomery et al., 2004; Terra and Montgomery, 1998) for 52 weeks) to relapse. We increased the recurrence rate by 30% for patients transitioning to a remission state from major depression without treatment (Hansen et al., 2008).

#### Suicide and Death

We assumed that depression-related suicides occur only among major depressive patients regardless of treatment status. We derived the transition probabilities for suicide from the modifications noted in (Valenstein et al., 2001) for age and gender-specific rates. The validity of suicide rates was ensured by comparing the suicide prevalence ratios from 2001 to 2018 (Center for Suicide Prevention, 2020; National Institute of Mental Health, 2019). We assumed that at least 50% of suicides were related to depression and 50-80% of older adults who die by suicide have been shown to have major depression (Center for Suicide Prevention, 2020).

Age-specific death from other causes was used (Arias et al., 2017), and we assumed that the major depressive patients had increased their risk of death based on other reasons by a ratio of 1.58 (Cuijpers et al., 2014).

#### Screening Tools

For the baseline model, the detection rate of depression in usual care settings increased by 50% (Valenstein et al., 2001) for major depression (baseline 45% vs. with screening 68%) and 23% (Valenstein et al., 2001) for minor depression (baseline 30% vs. 37% with screening).

#### Costs and Utilities

We conducted our analysis from the societal perspective, which took into account direct and indirect costs (Table 2). We assumed that patients who received treatment from primary care settings had 4 visits plus a follow-up visit in a year (Valenstein et al., 2001). Patients treated by mental health specialists had around 11 visits (Valenstein et al., 2001). 31% of patients received treatment from mental health specialists (26% self-refer and 5% referred by primary care physician) in a year (Valenstein et al., 2001). Additionally, 1% of patients have inpatient visits in a 12-month period with an average of 11.6 days of stay. On the other hand, the cost of minor depression was estimated as two-thirds of major depression (Cuijpers et al., 2007). We calculated the average treatment cost of partial remission (Israel, 2010) as $1095, based on the number of remaining symptoms. We assumed there is no direct or indirect cost to capture during full remission states, as for the healthy state.

**Table 2.** Cost and utilities for each health state.

| **COSTS** |  |  |  |
| --- | --- | --- | --- |
| **Variable** | **Description** | **Point Estimate (\$)** | **Reference** |
| Initial visit to primary care physician |  | 45.85 | (Valenstein et al., 2001) |
| Administration of screening cost |  | 4.88 | (Valenstein et al., 2001) |
| Screening cost |  | 50.73 | (Valenstein et al., 2001) |
| Treatment of depression in primary care settings | 4 visits and follow-up visits in a year | 398.72 | (Valenstein et al., 2001) |
| Treatment of depression by mental health specialist | 11 visit/year | 1020.14 | (Valenstein et al., 2001) |
| Medication cost (outpatient) |  | 677.16 | (Valenstein et al., 2001) |
| Hospitalization cost | Average length of stay 11.61 days | 11610 | (Valenstein et al., 2001) |
| Physician professional fees (hospital) | Daily inpatient physician visits (based on average length of stay) | 878.99 | (Valenstein et al., 2001) |
| Medication in hospital | Based on average length of stay | 21.83 | (Valenstein et al., 2001) |
| Indirect cost of major depression | Extrapolated from lost workdays of treated and untreated patients with major depression 1 year | 1600 for patients receiving treatment; 3360 for patients not receiving treatment | (Valenstein et al., 2001) |
| Minor depression with treatment | 2/3 of major depression cost | 2141 | (Cuijpers et al., 2007) |
| Minor depression without treatment | Indirect cost | 2101 | (Cuijpers et al., 2007) |
| Partial remission | on average 2-3 symptoms | 1095 | (Cuijpers et al., 2007), (Israel, 2010) |
| **UTILITIES** |  |  |  |
| **State** | **Description** | **Utilities** | **Reference** |
| Major depression | Same for with and without treatment | 0.63 | (Valenstein et al., 2001) |
| Minor depression, partial remission | Same for with and without treatment | 0.7 | (Valenstein et al., 2001) |
| Full remission | Same for full remission from minor and major depression | 0.89 | (Valenstein et al., 2001) |

The indirect cost was 20% lower in untreated patients ($1600 for treated vs. $3360 for untreated). The overall cost of major depression with treatment was estimated at $3087 vs. $3360 for major depression without treatment, which was similar to the estimates from (Cuijpers et al., 2007).

Utility estimates for all health states were obtained from the literature (Table 2). We assumed that depression with and without treatment states had the same utilities.

### Incremental Cost-effectiveness Ratio (ICER)

For all screening scenarii, costs and effects were compared to a situation without screening. Strategies were evaluated based on the incremental cost-effectiveness ratio (ICER), which was calculated as the incremental costs per incremental Quality-Adjusted Life Years (QALY) gained (Equation (1)). The scenario up to the willingness-to-pay (WTP) threshold of $50 000 per additional QALY gained was classified as optimal (Cohen and Reynolds, 2008).

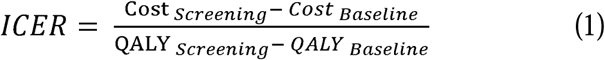

### Uncertainty Analysis

We performed a one-way sensitivity analysis on critical parameters (Valenstein et al., 2001). We obtained all the varying values of sensitivity analysis parameters from the literature (Supplementary Table S1). We used the medium and high values of incidence and prevalence (Eaton et al., 2007; Kessler et al., 2010) and matched two of these with using recall bias rates (Yildirim et al., 2021) for sensitivity analysis. Additionally, we tested sensitivity levels for other parameters such as remission rate from major depression treatment, the utility of major depression, cost of screening, treatment, and indirect costs.

We also performed Monte Carlo replications to report uncertainties with Bootstrapping-type analysis. We calculated the average incremental cost and QALY pairs to determine which proportion of ICER estimates lie below WPT, and 95% confidence intervals (CI) of ICERs.

## RESULTS

### Screening Impact on Outcome Measures

The mean duration in treatment was estimated from the simulation model as 1.26 years for major depression and 1.32 months for minor depression among females. The suicide rates in the baseline model were 5.11 for females and 18.08 for males per 100,000 people in a year. Annual screening could prevent 24.3 cases of suicide per 10,000,000 people in a year (Table 3). Annual screening also shortened the initiation of the treatment time by 5.3 (vs. 2.8 for 2-year screening and 1.1 for 5-year screening) months for females and 3.2 (vs. 1.7 for 2-year screening and 0.6 for 5-year screening) months for males. Annual screening increased the depression-free months up to 5.5 months per year. All the screening scenarii extended time spent in remission (11 days to 1.9 months for females and 7 to 29 days for males based on screening schedule).

**Table 3.**
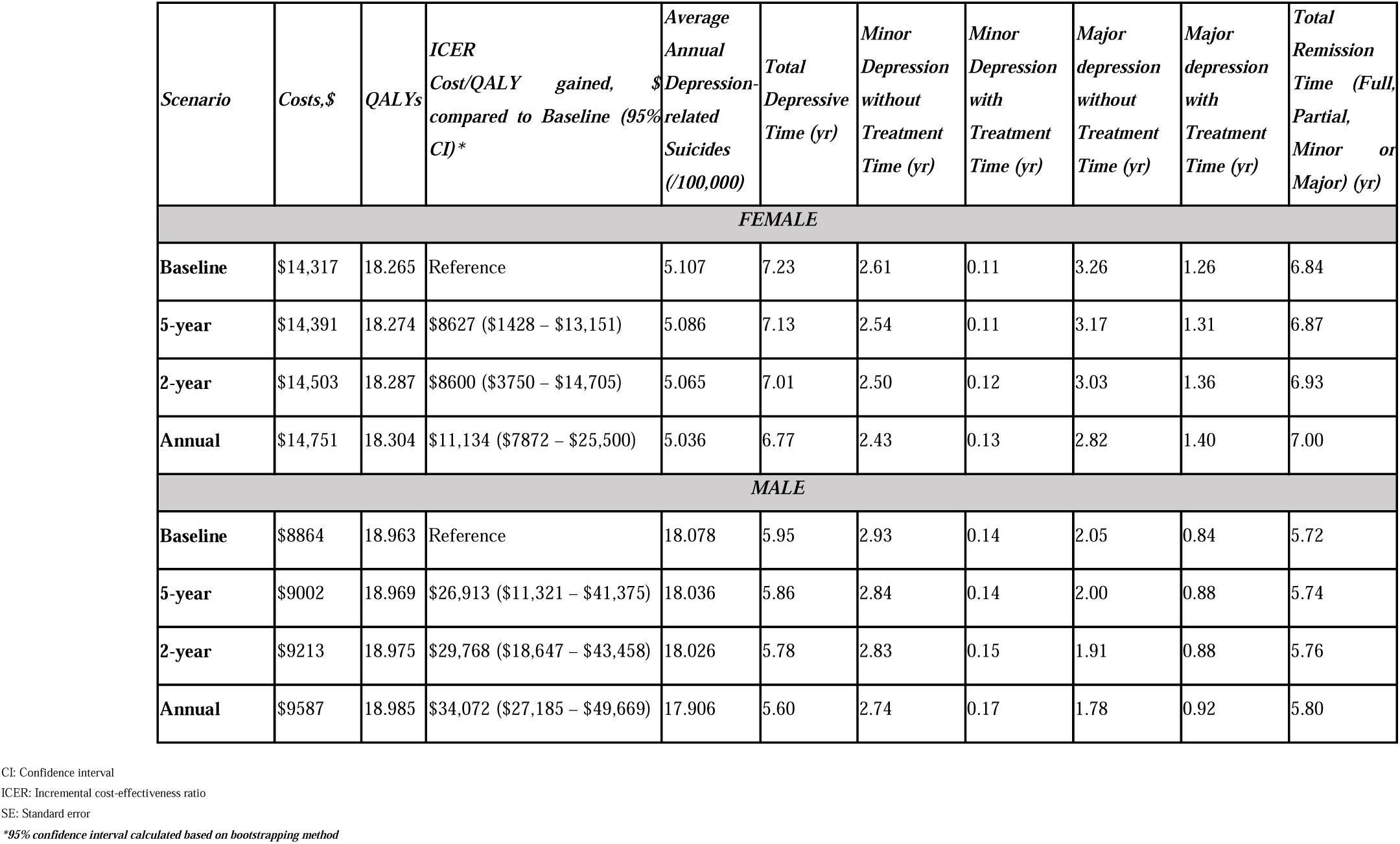
Cost-effectiveness measures of screening scenarios (average of 1000 replications)

Table 3 shows that, in the general population, all screening frequencies were cost-effective compared with the baseline. However, there exists a difference between the male and female populations based on cost-utility. Females had lower ICERs, and the annual screening had the highest ratio, with $11,134 cost per QALY gained. On the other hand, males had around 3 times higher ICER, with annual screening $34,072 cost per QALY gained.

### Uncertainty Analysis

#### One-way Sensitivity Analysis

In our baseline analysis, we observed that screening was cost-effective for both genders with low incidence and prevalence (conservative case). Consequently, the screening under medium and high incidence rates was cost-effective for females and males. For females, lower ICERs were observed compared to those for males.

Additional analysis on the other parameters such as the remission rate from major depression treatment, the utility of major depression, cost of screening, treatment, and indirect costs showed that female ICERs were not sensitive to any changes in these parameters. However, the ICERs of the male population were higher than $50,000 by using the upper limit of the screening (Figure 2).

#### Uncertainty among Monte-Carlo Simulations

Figure S2 (in supplementary) showed every single average incremental cost and QALY pairs for 1000 replications. For the female population, simulation results showed that all ICERs were less than the WTP threshold of $50,000. Additionally, less than 10 percent of simulation runs for males were greater or equal to the WTP threshold in every scenario (Figure S1).

### Age-Specific Analysis

Three different age groups, young (18-34), middle (35-64), and old (65+) were analyzed by evaluating differences in cost-utilities among these subgroups; see Figure S3 (in Supplementary).

All scenarii were cost-effective for each age group of the female population. For the oldest population, about 10% of the simulations have ICERs between $50,000 and $75,000 (please see Supplementary Figure S1).

Young and middle age groups of the male population had ICER values up to $32,090/QALY gained. On average, screening with any frequency was not cost-effective for the oldest male population.

## DISCUSSION

Annual screening of all adults 18 years old or older for depression would be $11,134 and $34,072 cost per QALY gained for females and males, respectively. In comparison, screening every 5-years resulted in $8627 cost per QALY gained for females and $26,892 cost per QALY gained for males. We found that the benefit of early detection and treatment results in the improvement of QALYs. For both genders, annual screening produced greater costs. The ICER decreased with increasing screening frequency for males, whereas it rose from 2-year to 5-year screening in the female population.

In the literature, the combined results from trials showed that depression screening increased the treatment rates between 2 to 50% (Pignone et al., 2002). In our model, patients spent an average of 3.65 years to initiate treatment for first-onset major depression. The World Health Organization ‘s (WHO ‘s) World Mental Health Survey showed that 35% of patients had 4 years of untreated time before initiating the treatment (Wang et al., 2005). We observed that screening decreased the detection time up to 5 months and increased the remission time up to 1.9 months.

Based on the general intuition, depression treatments could help to improve patient ‘s mood and depressive symptoms. However, some studies showed that 10% to 30% of patients experienced adverse outcomes from treatments (e.g., antidepressants) such as functional impairment, poor quality of life (Al-Harbi, 2012). Therefore, we chose conservative estimates for utilities. For example, we assumed that the utility of major depression with and without treatment was the same. Consequently, our results were not biased towards screening.

The age-specific analysis showed that screening of old populations had higher ICERs than the young and middle age groups. In the old male population, ICERs of all screening scenarii were below the WTP threshold of $50,000. The main reason for gender differences in the cost-effectiveness of screening was the varying incidence and prevalence of major depression. In literature, men had lower incidence rates than women (Eaton et al., 2007), although there is concern that these rates were not reflecting the truth (Smith et al., 2018). There were a couple of factors that may cause the underestimation of the rates for males. Firstly, the measurement bias was observed in data because of the inadequate attendance to surveys and lower primary care visits (Smith et al., 2018). Secondly, men were less likely than women to have symptoms of depression that fit standard measurement tools. They experienced more externalizing symptoms, such as aggression, violence, and substance abuse. Studies showed that including these alternative symptoms with traditional symptoms as diagnosis criteria had increased the male prevalence rates to have equal proportions with females (Martin et al., 2013). We evaluated the alternative incidence, prevalence, and screening scenarii for males with the same frequency as females. Our results are summarized in the Supplementary file.

We evaluated the external consistency of our model. By running the open network model for a longer time, where new individuals were introduced into the system at the age of 18 with the mortality rate, we observed that our initial distribution was equal to steady-state distribution. This was the case when transient behavior was not observed in the system. To obtain a consistent system, we adjusted the lifetime prevalence of major depression using reported recall bias rates (Yildirim et al., 2021).

Interventions (e.g., training primary care physicians to better identify patients with suicidal thoughts) that have an immediate cost but have gains observed during many years were less cost-effective under discounting. For interventions like screening, the costs and health benefits follow the same time pattern so are all affected equally by the discount factor, and the relative ICER comparisons between policies are unchanged, regardless of the discounting (Jamison et al., 2006). We ran additional computational results (similar to Table 3) with a discount factor. In each case, the ICER that we found fell within the 95% confidence interval on the ICER currently in Table 3. We also find that the relative ICER comparisons between policies continue to hold, such as the cost-effectiveness of the screening for the general adult population, 2-year screening ICER is less (more) than 5-year screening ICER for females (males). Therefore, we did not include the discount rate of utilities and costs in the body of the paper.

This paper evaluated the general U.S. population during the non-crises period (e.g., economic problems, or pandemics). Recent studies showed that during COVID-19, incidence and prevalence were reported three times higher for moderate and severe depression (Ettman et al., 2020). We further included the prevalence rate changes between 2020 and 2021 in our analysis. Our results indicated that the depression screening is cost-saving for females at 2 and 5-year screening frequencies and around $11,000 to $15,000 ICERs for males based on the screening interval between 2020 to 2040 (please see Supplementary file).

## LIMITATIONS

The cost-effectiveness of screening may be enhanced by targeting groups with a higher incidence of depression based on ethnicity, comorbidities, or poverty level. However, we analyzed screening scenarii that are valid for the general adult population. Because of incidence, disease progression, and suicide rate difference between females and males, we only considered the gender-specific model. We also extended our analysis for different age groups.

Our model did not consider the differences among the screening tools; we used average test sensitivity for 9 standard screening instruments. We assumed that the screening frequency was not changing at any time during the screening scenario. In our calculations, false-positive cases were not taken into account.

We did not consider the severity of major depression; however, we included the details about elevated risk based on age, gender, history of depression (major or minor), number of previous episodes, remaining symptoms for onset depression, treatment status, type of treatment and time spent without treatment states. We did not exclusively include medication drop-out rates (transition from treatment to without treatment) in our analysis. However, the treatment rates obtained from the literature implicitly include the response of adherent and non-adherent patients who are not fully complied the clinical treatment guidelines.

Like Valenstein ‘s model, (Valenstein et al., 2001) we assumed that suicide might happen when the patient had active major depression. Patients did not have direct and indirect costs (e.g., productivity loss) when they were in full remission; however, staying there for a long time decrease the risk of relapses. But distinct from their work, we used more detailed remission states, e.g., partial and full remission from different types of depression. We did not consider the medication costs that arose when patients were in remission. In the annual transition model, patients spent at least one year in treatment when they transition to treatment states, which is more than suggested acute therapy (6-12 weeks) and continuation therapy (4-8 months). We assumed that patients in partial remission with residual symptoms had received maintenance treatment.

## CONCLUSIONS

Depression is a common health condition that affects an individual ‘s everyday life. Considering the high lifetime prevalence and recurrence rates of depression, detection and prevention efforts play an essential role. Furthermore, the increasing trend in suicide rates becomes an emerging public health problem. We conclude that routine screening is cost-effective for all age groups of females and young and middle-aged males. Male population results are sensitive to the higher costs of screening, which indicates that if the screening cost is 44% higher than the average cost, the screening of the male population is not cost-effective. Our analysis combines the expected benefits and costs of screening and assesses the effectiveness of screening scenarii. Screening can be one of the cornerstones to support required care. Our baseline model could be used to evaluate the potential consequences of medication strategies or alternative intervention scenarii.

## Data Availability

All data used in the study is publicly available as described in the paper.

## Acknowledgements

The authors are grateful to the Editors and anonymous reviewers for their thoughtful, constructive comments.

