## Supplementary file for "THE COST-EFFECTIVENESS OF DEPRESSION SCREENING FOR THE GENERAL ADULT POPULATION"

This supplementary material file has been created by the authors to give readers additional information about the research article with the above title.

**Appendix 1.** Additional table and figure

**Figure S1.** Uncertainty for age groups

**Table S1.** Sensitivity Analysis Parameters

**Figure S2.** Incremental cost-effectiveness plane

**Figure S3.** Age-specific incremental cost-effectiveness ratios (ICERs)

**Appendix 2.** ICER estimates with a discount rate

**Table S2.** ICERs with a discount rate of 3%

**Appendix 3.** Case of COVID-19

**Table S3.** Incidence and prevalence updates

**Table S4.** Results for Scenarios 1 and 2

**Appendix 4.** Evaluation of Gender Differences (Male)

**Figure S4**. ICER of Screening vs. Baseline

**References**

***Appendix 1.*** Additional table and figure.

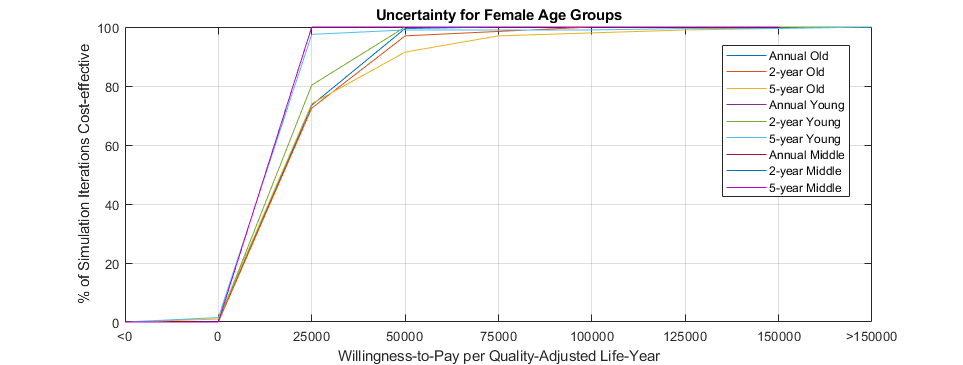
***Figure S1.*** *Uncertainty for age groups*

Female

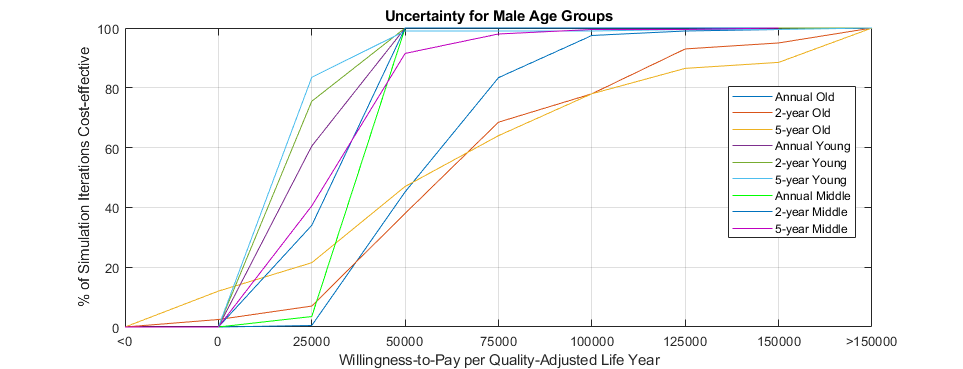

Male

***Table S1.*** *Sensitivity Analysis Parameters*

| **Definition** | **Probability, Ratio or Cost** | **Age Group** | **Gender-specific** | **Reference** |
| --- | --- | --- | --- | --- |
| ***Major Depression Prevalence % and Incidence*** | | | | |
| Without treatment [medium-high] | **Female** [11.85-12.95], [9.85-10.76], [7.93-8.66], [3.22-3.52], **Male** [6.01-7.21], [6.18-7.41], [5.09-6.11], [1-1.2] | 18-34, 35-49, 50-64, 65+ | Yes | Calculated (Kessler et al., 2010), (Yildirim et al., 2021) |
| With treatment [medium-high] | **Female** [6.94-7.59], [5.77-6.3], [4.65-5.08], [1.89-2.06] **Male** [3.28-3.93], [3.37-4.04], [2.78-3.33], [0.5-0.66] | 18-34, 35-49, 50-64, 65+ | Yes | Calculated (Kessler et al., 2010), (Yildirim et al., 2021) |
| Full remission [medium-high] | **Female** [9.96-10.88], [23.15-27.2], [25.62-30.69], [30.81-36.93] **Male** [8.84-9.34], [15.68-19.23], [16.52-21.13], [21.02-27.21] | 18-34, 35-49, 50-64, 65+ | Yes | Calculated (Kessler et al., 2010), (Yildirim et al., 2021) |
| Incidence [medium-high] | **Female** [0.0045-0.0088], [0.0065-0.0092], [0.0024-0.0046], [0.001-0.0028], **Male** [0.0015-0.0052], [0.0038-0.0065],[0.0009-0.0032], [0.0007-0.0037] | 18-29, 30-44, 45-64, 65+ | Yes | (Eaton et al., 2007) |
| ***Screening Cost*** | | | | |
| Initial visit to primary care physician [low-high] | [21.88-72.24] | No | No | (Valenstein et al., 2001) |
| Administration of screening cost ($) [low-high] | [3-10] | No | No | (Valenstein et al., 2001) |
| ***Major Depression Medical Costs*** | | | | |
| Treatment of depression in primary care settings [low-high] | [175-626.6] | No | No | (Valenstein et al., 2001) |
| Treatment of depression by mental health specialist [low-high] | [412.6-1327.6] | No | No | (Valenstein et al., 2001) |
| Medication cost ($) (outpatient) [low-high] | [513-863] | No | No | (Valenstein et al., 2001) |
| Hospitalization cost ($) [low-high] | [5600-16800] | No | No | (Valenstein et al., 2001) |
| Physician professional fees (hospital) [low-high] | [529-1059] | No | No | (Valenstein et al., 2001) |
| Medication in hospital [low-high] | [16.57-27.86] | No | No | (Valenstein et al., 2001) |
| ***Major Depression Indirect Costs*** | | | | |
| Patients receiving treatment, patients not receiving treatment [low-high] | [800,2400-1120,4480] | No | No | (Valenstein et al., 2001) |
| ***Major Depression Remission*** | | | | |
| In usual care to full remission [low-high] (25% difference from base) | [0.377-0.629] | No | No | (Judd et al., 1998) |
| ***Utility*** | | | | |
| Major depression [low-high] | [0.55-0.68] | No | No | (Valenstein et al., 2001) |

***Figure S2.*** *Incremental cost-effectiveness plane*

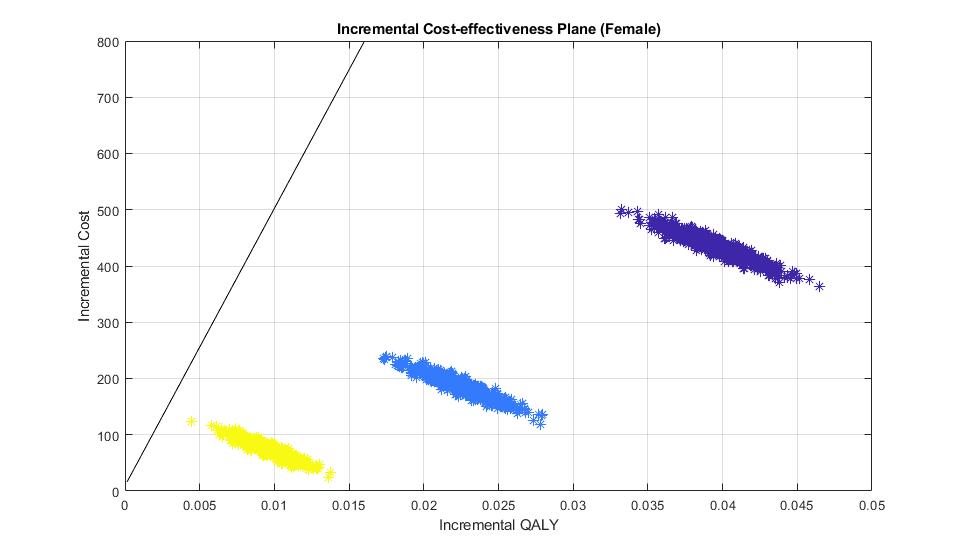

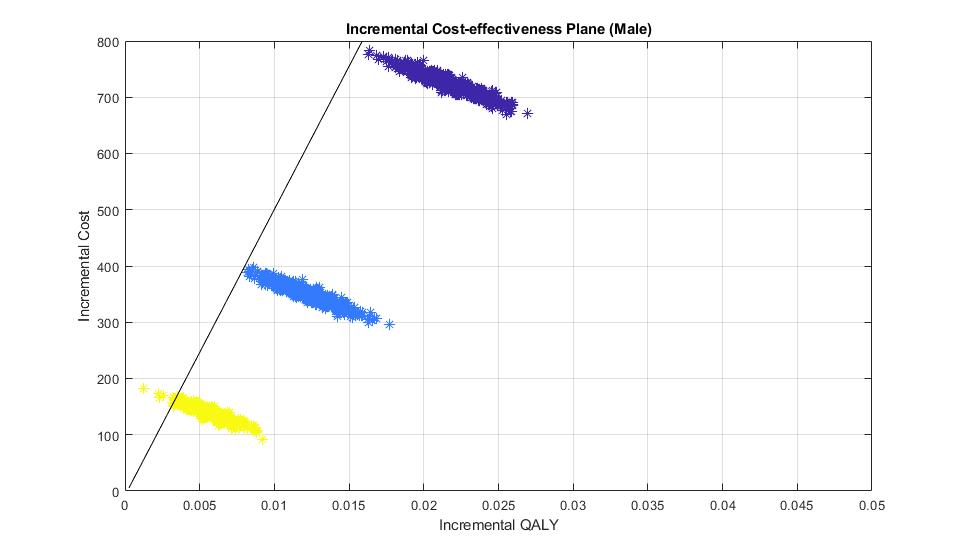

Female

Male

Annual

2-year

5-year

Willingness to pay threshold $50,000.

Willingness to pay threshold $50,000.

***Figure S3.*** *Age-specific incremental cost-effectiveness ratios (ICERs)*

**Effectiveness (QALYs)**

Young (18-34)

Middle (35-64)

Annual

2-year

5-year

Baseline

Old (65+)

$7K

$7.5K

$8K

$8.5K

$9K

$9.5K

$10.5K

$11K

$11.5K

$12K

$12.5K

$13K

$13.5K

$14K

$14.5K

$15K

17.8

18.0

18.2

18.4

18.6

18.8

19.0

19.2

19.4

19.6

$9155/QALY

$6276/QALY

$4783/QALY

$8676/QALY

$6705/QALY

$6474/QALY

$20,326/QALY

$18,834/QALY

$15,501/QALY

$23,738/QALY

$20,410/QALY

$14,942/QALY

$32,090/QALY

$26,792/QALY

$26,802/QALY

$51,094/QALY

$56,921/QALY

$64,845/QALY

$15.5K

**Costs**

Female

Male

***Appendix 2*.** ICER estimates with a discount rate

During our analysis, we did not include a discount rate in ICER estimates. In the below analysis, we added a 3% discount rate for cost and QALYs and reran the models to estimate the ICERs as reported in Table 3. The values found are within the 95% confidence interval as reported in Table 3.

***Table S2.*** ICERs with a discount rate

| **Scenario** | **Costs, $** | **QALYs** | **ICER**  **Cost/QALY gained, $ compared to Baseline** |
| --- | --- | --- | --- |
| **FEMALE** | | | |
| **Baseline** | $10,651 | 13.840 | Reference |
| **5-year** | $10,711 | 13.846 | $9,571 |
| **2-year** | $10,800 | 13.856 | $9,490 |
| **Annual** | $11,001 | 13.868 | $12,429 |
| **MALE** | | | |
| **Baseline** | $6,634 | 14.358 | Reference |
| **5-year** | $6,742 | 14.362 | $29,162 |
| **2-year** | $6,902 | 14.367 | $30,862 |
| **Annual** | $7,193 | 14.373 | $37,267 |

***Appendix 3*.** Case of COVID-19

i) **Covid-19 and model updates: incidence and prevalence**

Mental health problems are underdiagnosed and undertreated in general circumstances. Thus, the identification of patients requires an additional effort. As shown in the literature, most individuals had experienced stress, anxiety disorders, or depression during and after public health emergencies. (Cheng et al., 2004) Therefore, it is vital to consider individuals and factors they encounter during the COVID-19 outbreak and target individuals who need mental health interventions.

Major depression prevalence was estimated as three times higher during the outbreak, and minor depression prevalence was 50% higher than the 2019 prevalence rates. (Ettman et al., 2020) As shown in Table S2, we updated prevalence rates according to this. The new incidence rate was calculated to match the prevalence rates in the same way as Yildirim et al. (Yildirim et al., 2021)

***Table S3.*** Incidence and prevalence updates

|  | Annual Prevalence | | | Annual Incidence | | |
| --- | --- | --- | --- | --- | --- | --- |
|  | Before Covid-19* | During Covid-19 | Multiplier | Before Covid-19 | During Covid-19 | Multiplier |
| Female | 0.102^a^ | 0.3^b^ | 3 | 0.0029^c^ | 0.0348^d^ | 12 |
| Male | 0.062^a^ | 0.18^b^ | 3 | 0.0013^c^ | 0.013^d^ | 10 |
| References | (Kessler et al., 2010) | (Ettman et al., 2020) | (Ettman et al., 2020) | (Eaton et al., 2007) | Calculated (Eaton et al., 2007; Yildirim et al., 2021) | Calculated |

*Lower Bounds

ii) **Simulation Scenarios**

In scenarios 1 and 2, we simulated the model for annual, 2-year, and 5-year screening scenarios. All the parameters and probabilities were the same as the model provided in the manuscript, except incidence and prevalence.

a. Scenario 1:

In this scenario, we evaluated March 2020 to March 2025 and assumed that COVID-19 incidence and prevalence are lasting 5 years. We further calculated age-specific incidence and prevalence rates using a multiplier in Table S2.

b. Scenario 2:

With this scenario, we continued to evaluate the 20-year time horizon. We assumed that simulation starts in 2020 with the incidence and prevalence during COVID-19, provided in Table S2. We quantified the annual incidence as it was dropping by 20% compared to the previous year. Between 2025 and 2040, the incidence will be the same as the average incidence before COVID-19.

iii) **Results**

Both scenarios showed that screening was cost-effective for females and males. The results were summarized in Table S3. ICERs were varying based on the simulation horizon, screening schedules, and gender.

***Table S4.*** Results for Scenarios 1 and 2

| Gender | Screening | Costs, $ | QALYs | ICER | Average Annual Depression-related Suicides (/100000) |
| --- | --- | --- | --- | --- | --- |
| Scenario 1 | | | | | |
| Female | Baseline | 5675 | 4.020 | Reference | 5.922 |
|  | Annual | 5789 | 4.026 | 17590 | 5.628 |
|  | 2-year | 5730 | 4.024 | 16117 | 5.637 |
|  | 5-year | 5697 | 4.022 | 14951 | 5.841 |
| Male | Baseline | 3721 | 4.292 | Reference | 28.033 |
|  | Annual | 3897 | 4.295 | 48907 | 27.640 |
|  | 2-year | 3809 | 4.294 | 48114 | 27.756 |
|  | 5-year | 3756 | 4.293 | 46897 | 27.976 |

| Scenario2 | | | | | |
| --- | --- | --- | --- | --- | --- |
| Female | Baseline | 22325 | 16.958 | Reference | 8.368 |
|  | Annual | 22340 | 17.024 | 238 | 8.297 |
|  | 2-year | 22277 | 16.995 | -1290 | 8.493 |
|  | 5-year | 22305 | 16.973 | -1306 | 8.476 |
| Male | Baseline | 13498 | 18.107 | Reference | 30.544 |
|  | Annual | 14021 | 18.141 | 15427 | 30.061 |
|  | 2-year | 13734 | 18.126 | 12456 | 30.192 |
|  | 5-year | 13588 | 18.115 | 11008 | 30.378 |

iv) **Conclusions**

To prevent the unexpected outcomes of untreated depression, such as chronic depression, longer medication usage, and suicide. It is essential to intervene early and detect the cases before disease progression. It is known that untreated depression increases the risk of other conditions (e.g., use of illicit drugs). (Li et al., 2020) We conclude that both genders' screening in a 5-year or 20-year time horizon will be cost-effective. Considering incidence and prevalence rate increases due to the COVID-19, screening scenarios for 20-years will be more cost-effective than before COVID-19 estimates.

***Appendix 3.*** Evaluation of Gender Differences (Male)

We further evaluated the case that male population incidence and prevalence were underestimated in the literature. We tested the following hypothesis.

Male incidence and prevalence equal that of females:

In literature, including alternative (aggression, violence, etc.) and classic depression symptoms found that men and women had been diagnosed with depression in equal proportions. (Martin et al., 2013) Adjustment on male incidence and prevalence rates shows that the average population was cost-effective with all screening schedules (Figure S4). Old males were also cost effective (<50,000 cost/QALY) in 95% of simulation replications.

***Figure S4***. ICER of Screening vs. Baseline

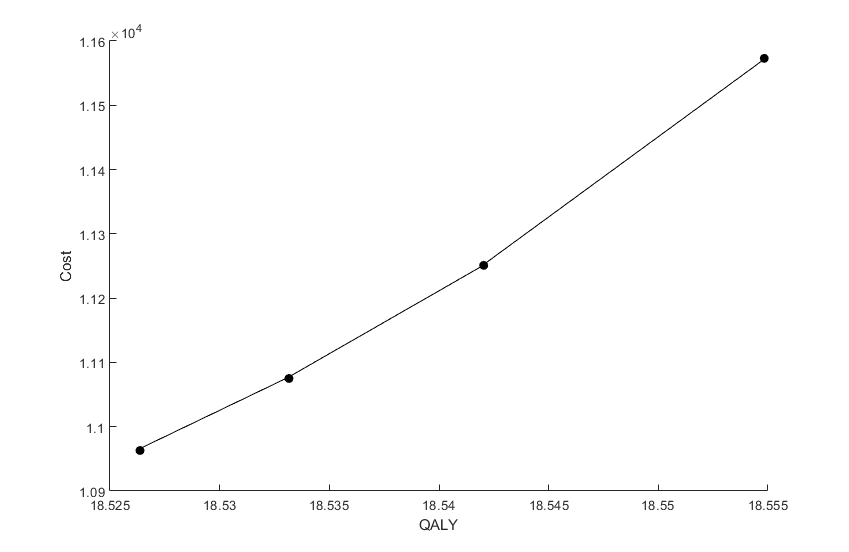

Baseline

5-year

2-year

Annual

$21,576/QALY

$18,726/QALY

$17,423/QALY
